# Causes of Death and Comorbidities in Patients with COVID-19

**DOI:** 10.1101/2020.06.15.20131540

**Authors:** Sefer Elezkurtaj, Selina Greuel, Jana Ihlow, Edward Michaelis, Philip Bischoff, Catarina Alisa Kunze, Bruno Valentin Sinn, Manuela Gerhold, Kathrin Hauptmann, Barbara Ingold-Heppner, Florian Miller, Hermann Herbst, Victor Max Corman, Hubert Martin, Helena Radbruch, Frank L. Heppner, David Horst

## Abstract

Infection by the new corona virus strain SARS-CoV-2 and its related syndrome COVID-19 has caused several hundreds of thousands of deaths worldwide. Patients of higher age and with preexisting chronic health conditions are at an increased risk of fatal disease outcome. However, detailed information on causes of death and the contribution of comorbidities to death yet is missing. Here, we report autopsy findings on causes of death and comorbidities of 26 decedents that had clinically presented with severe COVID-19. We found that septic shock and multi organ failure was the most common immediate cause of death, often due to suppurative pulmonary infection. Respiratory failure due to diffuse alveolar damage presented as the most immediate cause of death in fewer cases. Several comorbidities, such as hypertension, ischemic heart disease, and obesity were present in the vast majority of patients. Our findings reveal that causes of death were directly related to COVID-19 in the majority of decedents, while they appear not to be an immediate result of preexisting health conditions and comorbidities. We therefore suggest that the majority of patients had died of COVID-19 with only contributory implications of preexisting health conditions to the mechanism of death.

## INTRODUCTION

More than 7.9 million confirmed cases of coronavirus disease 2019 (COVID-19) and more than 434 thousand associated deaths have been counted around the globe by June 15, 2020^1^. COVID-19 is caused by infection with severe acute respiratory syndrome coronavirus 2 (SARS-CoV-2), a novel and highly contagious coronavirus strain that mostly spreads through respiratory droplets and that has first been identified in Wuhan, China^2^.

Some SARS-CoV-2 infections are asymptomatic while most cause mild to moderate illness with respiratory and flu-like symptoms, including fever, chills, cough and sore throat^3,4^. However, a significant number of patients with COVID-19 develops critical illness and requires intensive care with mechanical ventilation or extracorporeal membrane oxygenation^5,6^. Especially in these cases, the disease may ultimately be fatal^7^. While raw numbers of deaths suggest overall COVID-19 case-fatality rates of more than 5 %^1^, infection-fatality rates probably are lower and may range around 0.3-0.5 %^8^.

The risk of death from COVID-19 strongly depends on age and previous health conditions. Older patients and those with chronic comorbidities, such as cardiovascular disease, hypertension, diabetes, and pulmonary disease, are much more prone to critical and fatal disease outcomes^4,9^. These associations may contribute to an uncertainty to what extent COVID-19 or preexisting health conditions determined the time of a patient’s death. Detailed information on the causality and mechanism of death, as well as the spectrum of comorbidities in cases with fatal outcome that will allow accurate assessment of the hazardous nature of COVID-19 yet is missing.

Autopsies are the gold standard for the analysis of medical conditions and causes of death. Few reports described damage to several organs due to COVID-19 on the morphological level. However, while one study reported fatal pulmonary thromboembolism in few cases^10^, systematic information on immediate and underlying causes of death are scarce. Here, we present data on clinical and autoptic causes of death and comorbidities of 26 patients that had died after SARS-CoV-2 infection and COVID-19 in Berlin, Germany. Our findings reveal that causes of death were directly related to COVID-19 in most cases and not an immediate consequence of preexisting health conditions and comorbidities, i.e. these patients – despite often suffering from severe health conditions – would not have died in the absence of a SARS-CoV-2 infection at the given time point.

## METHODS

### Study design

26 cases of patients that had died after COVID-19 disease were included. In all cases, SARS-CoV-2 infection was confirmed by PCR testing of material from nasal and pharyngeal swabs. In 24 cases, patients had been treated at Charité – Universitätsmedizin Berlin, while two were referred from Vivantes – Netzwerk für Gesundheit GmbH and DRK Kliniken Berlin. Autopsies were performed on the legal basis of §1 SRegG BE of the autopsy act of Berlin and §25(4) of the German Infection Protection Act. This study was approved by the Ethics Committee of the Charité (EA 1/144/13 and EA2/066/20) as well as by the Charité-BIH COVID-19 research board and was in compliance with the Declaration of Helsinki. Clinical information on comorbidities and pre-existing conditions was obtained from patient files and clinical death certificates.

### Autopsy procedure

External examination, complete autopsy and tissue sampling were performed in 26 patients with COVID-19 according to the traditional technique by opening all luminal structures and lamellar incisions of all parenchymatous organs. According to recent recommendations for the performance of autopsies in cases of suspected COVID-19, safety precautions including FFP2-masks, protective suits and cut resistant gloves were applied for all autopsies^11^. For histopathology, representative tissue samples of all organs were fixed in 4 % buffered formalin, dehydrated, paraffin embedded and sectioned with a thickness of 4 µm. Paraffin sections were stained with hematoxylin and eosin (HE), periodic acid Schiff’s reaction (PAS), Van Gieson’s elastic stain, Prussian blue stain and Kongo-red stain. At least two pathologists examined all slides by light microscopy.

### Statistical analysis

Data collection and statistical analysis were done with IBM SPSS Statistics, Version 23 (IBM, NY, USA). Age at death was presented as median with interquartile range (IQR) to account for deviations from normal distribution. Categorical variables were summarized as counts and percentages. Median time to death was analyzed using the Kaplan-Meier method.

## RESULTS

### Clinical presentation and causes of death in COVID-19 decedents

We analyzed 26 cases of patients that died after COVID-19 disease and that were autopsied at the Institute of Pathology of the Charité university hospital in Berlin. In all cases, SARS-CoV-2 infection was confirmed by PCR testing. Of the decedents, 17 were male and 9 were female. One of the decedents was of Black ethnicity, while 25 were Caucasians. The median age at death was 70 years (IQR: 61.8 - 78.3, range: 30 to 92 years), and the time from onset of COVID-19 symptoms to death ranged from 5 to 51 days, with a median of 26 days. Additional clinical information on the course of the disease was available in all cases. Prior to death, all patients had presented with COVID-19 related lung disease. Signs of respiratory failure were most prevalent with 88.5 %, while in 57.7 % patients had signs of bacterial pneumonia. Furthermore, pulmonary thromboembolism was reported in 23.1 % of the cases with clinical evidence of deep venous thrombosis in two patients (7.7 %). Due to the severity of lung damage, patient care warranted invasive ventilation in 76.9 %, prone positioning in 53.8 %, and extracorporeal membrane oxygenation in 30.8 % (Table 1). Aside from lung involvement, acute renal failure was the second most prevalent organ failure and hemodialysis was necessary in 69.2 % of the patients. Furthermore, half of the patients presented with multi-organ failure, while acute liver failure was reported in 30.8 %. These findings indicated severe and complex courses of COVID-19 in these patients. An overview of the clinical characteristics is given in Table 1.

**Table 1:**
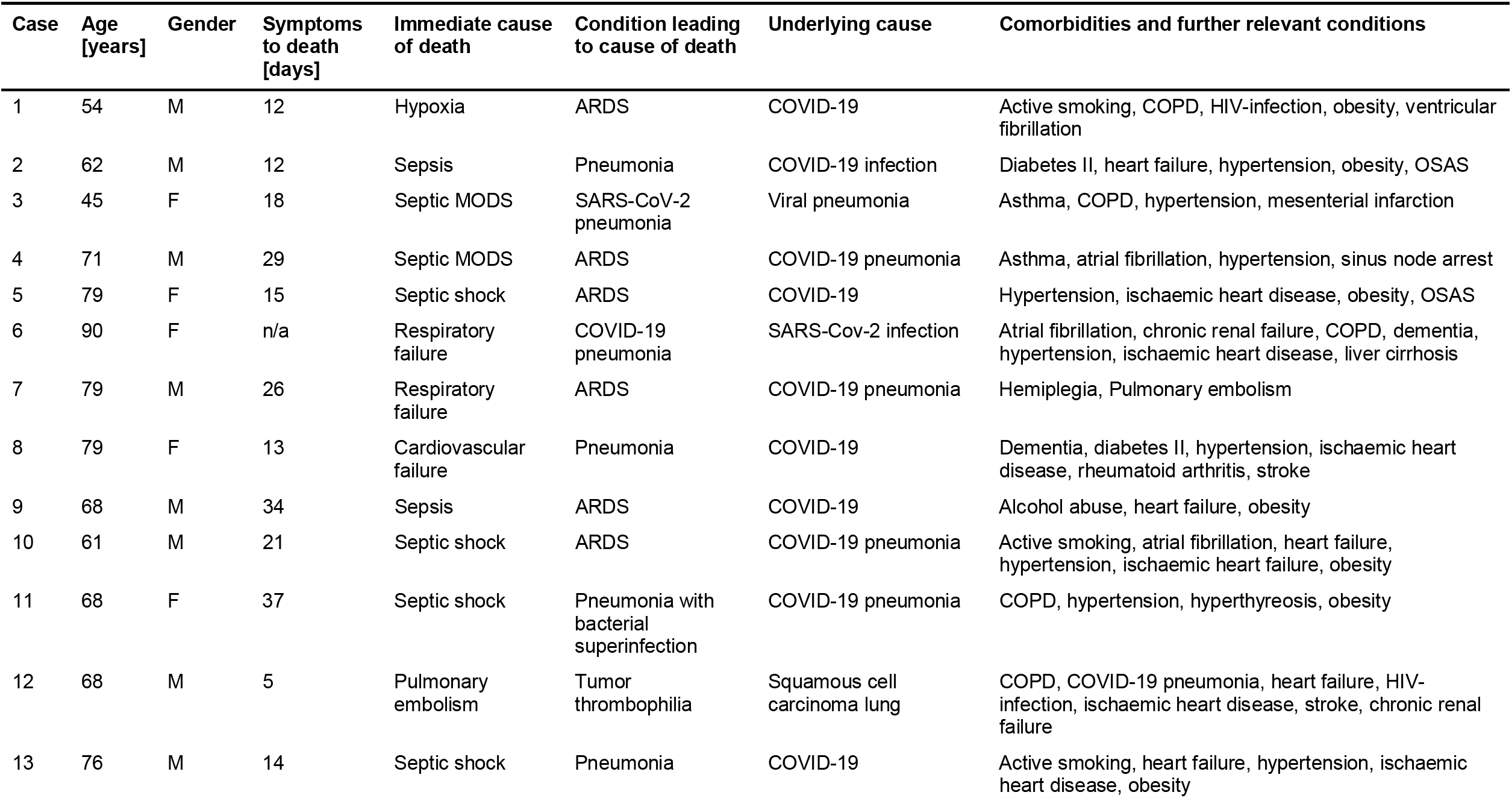
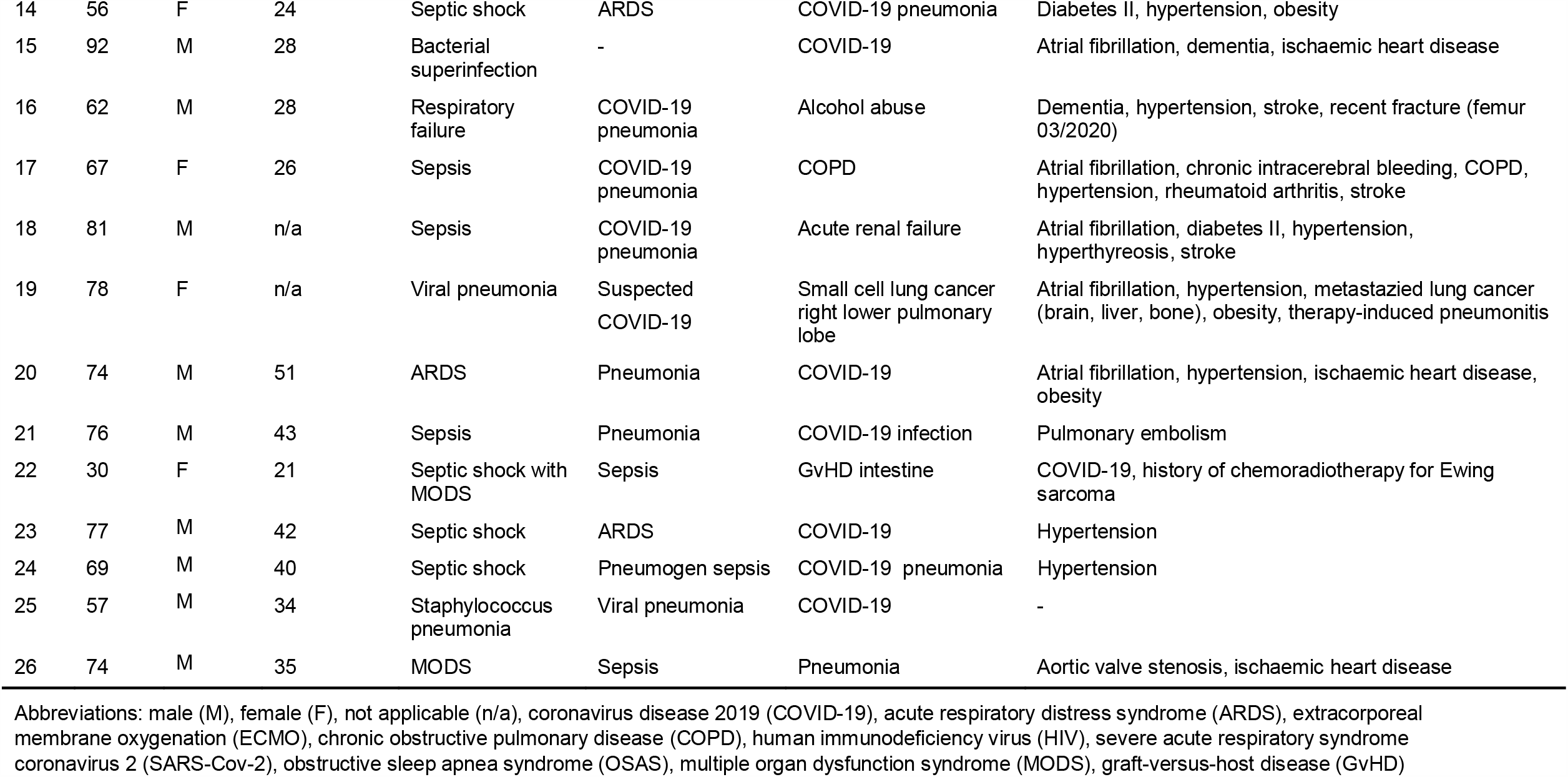
Clinical causes of death and documented comorbidities in hospitalized patients with COVID-19

To learn about clinical causes of death, we assessed legal death certificates of the 26 decedents. Most frequent immediate causes of death, documented in 19 cases (73.1 %), were infection related, and included sepsis, septic shock, or sepsis-related multi-organ failure in 16 cases (61.5 %), bacterial infections in two cases (7.7 %), and viral pneumonia in one case (3.8 %). Second most common were respiration-related causes of death, documented as respiratory insufficiency, hypoxia, or acute respiratory distress syndrome (ARDS) in four cases (15.4 %). Further individual immediate causes of death were pulmonary embolism and cardiovascular failure (3.8 % each).

In addition, clinical death certificates provided information on conditions leading to immediate causes of death and death-related underlying disease (Table 1). Here, pulmonary disease in 22 decedents (84.6 %) was the most frequently documented condition leading to cause of death, which included pneumonia or viral pneumonia in 14 cases (53.8 %), and ARDS in eight cases (30.8 %). Importantly, when jointly considering conditions leading to cause of death and underlying disease, COVID-19, confirmed or suspected SARS-CoV-2 infection was documented in a total of 23 cases (88.5 %), and thus in the vast majority of deceased patients. Other underlying diseases that were deemed clinically relevant for death included cancer in two cases (7.7 %), and individual cases of alcohol abuse, renal failure, COPD, or GvHD. Collectively, these data indicated that, from a clinical perspective, COVID-19 and infection-related disease were major contributors to patients’ death in the majority of cases.

### Clinical information on comorbidities

Clinical records also contained information on chronic comorbidities and further relevant health conditions. The median number of chronic comorbidities in these cases was four, and ranged from three to eight (Table 1). Arterial hypertension was the most prevalent chronic condition in the decedents (65.4 %), followed by obesity (38.5 %), chronic ischemic heart disease (34.6 %), atrial fibrillation (26.9 %), and chronic obstructive pulmonary disease (23.1 %). Vascular conditions were specified as atherosclerosis (7.7 %) and cerebrovascular disease (15.4 %). Of all patients, 15.4 % had diabetes type II, and chronic renal failure was noticed in 11.5 % of decedents. Active or non-active nicotine abuse was noted in 5 patients (19.2 %) and alcohol abuse in 3 patients (11.5 %). Further details are available from Table 1. These data suggested severe chronic comorbidities and health conditions in the majority of patients that had died after COVID-19.

### Causes of death determined at autopsy in decedents with COVID-19

In order to investigate causes of death directly, we performed full body autopsies including histopathological workup on all 26 decedents. Based on assessment of pathological disease mechanisms and referring to clinical documentations of death, we defined immediate causes of death, conditions leading to cause of death, and underlying causes (Table 2). As the most common immediate cause of death, we found septic shock and/or multi-organ failure in 8 patients (30.8 %), followed by suppurative pulmonary infections in five patients (19.2 %), including bacterial pneumonia with or without abscess formation, as well as infarct necrosis with signs of purulent bacterial superinfection. Right ventricular congestive heart failure or decompensation as immediate cause of death was present in four patients (15.4 %). In five patients (19.2 %), respiratory failure or diffuse alveolar damage was the immediate cause of death, with severe lung damage implicating highly restricted gas exchange. Four more cases presented with either deadly pulmonary thromboembolism, severe bronchial aspiration, gastrointestinal bleeding, or signs of left ventricular heart failure (3.8 % each).

**Table 2:**
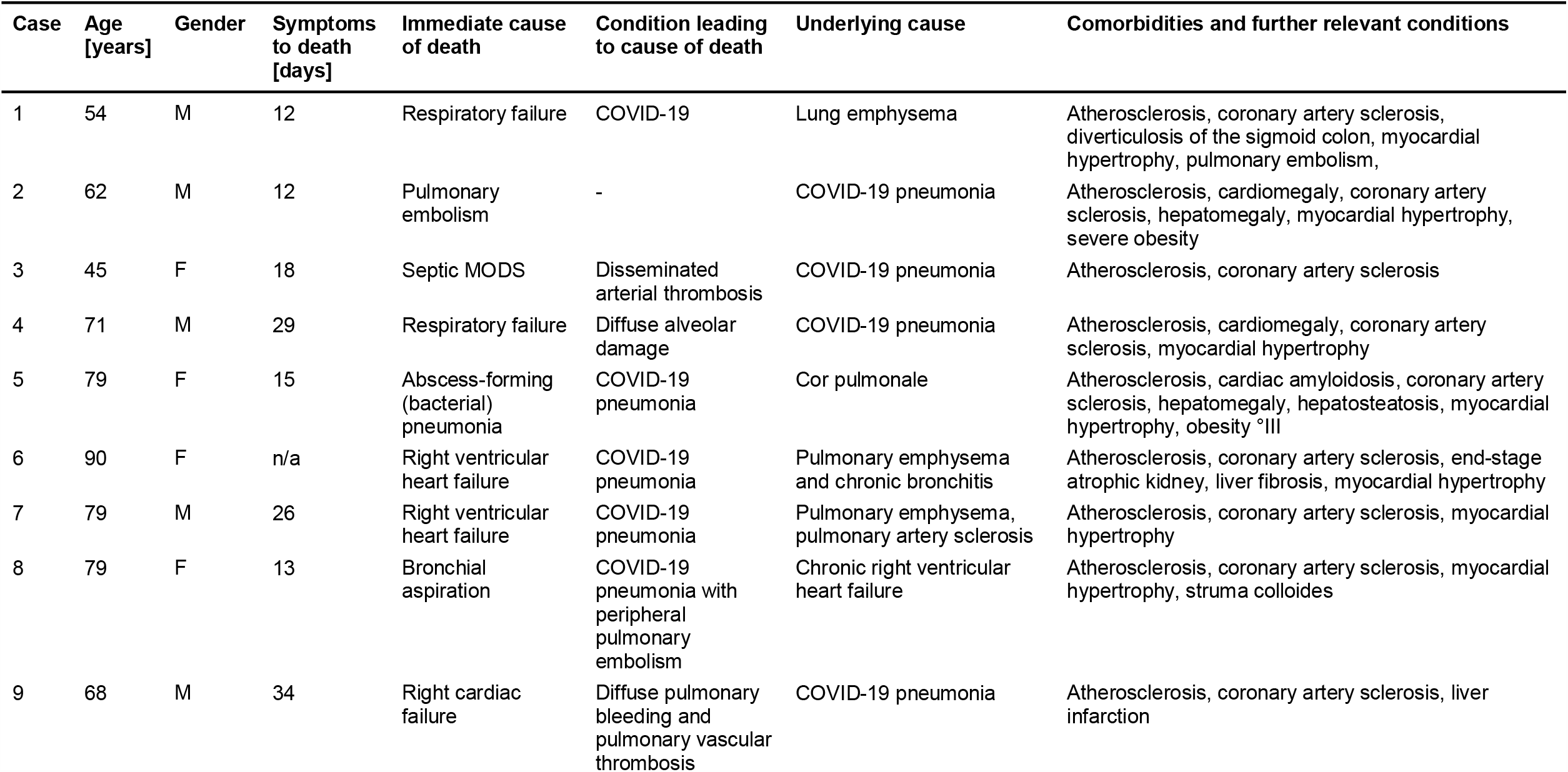
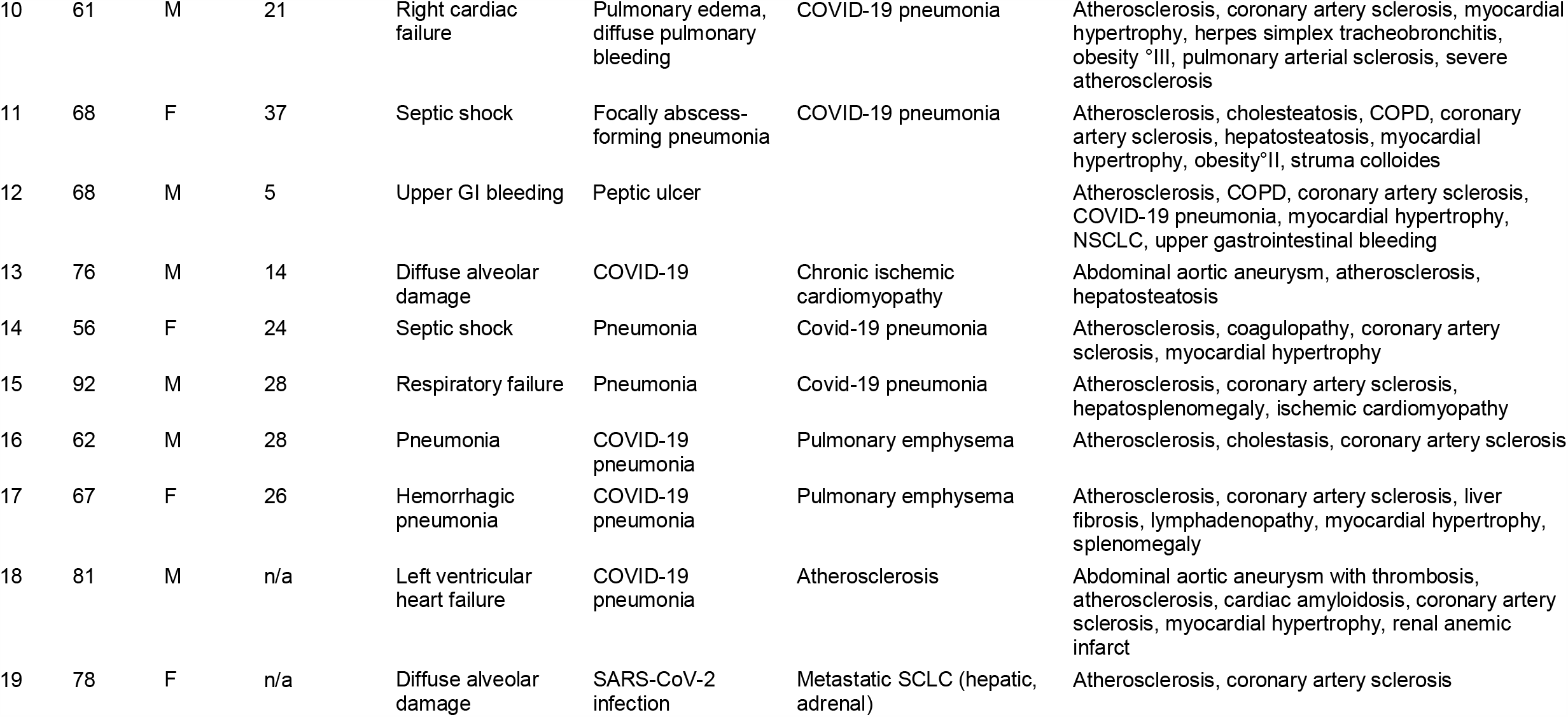
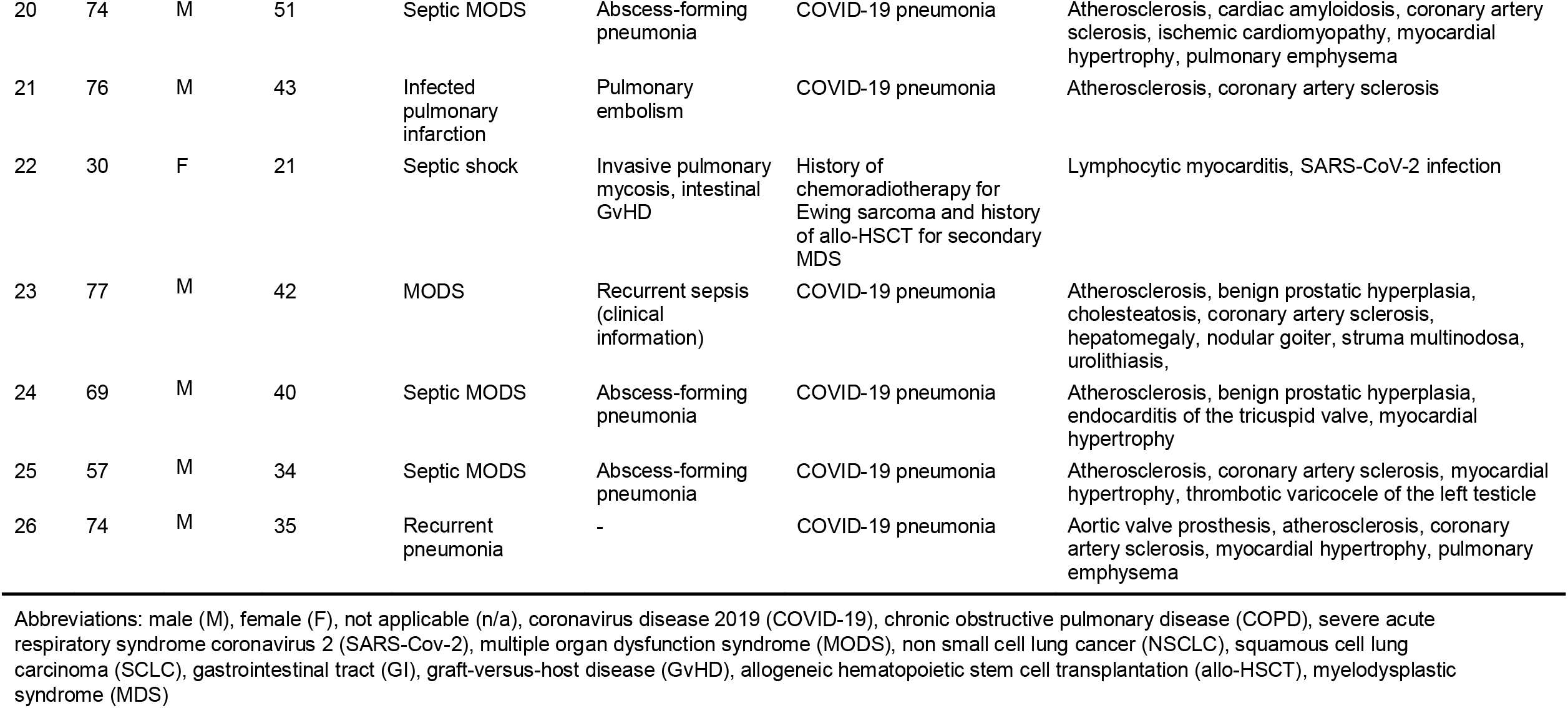
Causes of death and comorbidities in Patients with COVID-19 as determined by autopsy

We then determined conditions leading to these immediate causes of death (Table 2). We found that COVID-19 or SARS-CoV-2 infection most prevalently preceded the immediate cause of death in ten cases (38.5 %), followed by bacterial pneumonia with or without abscess formation in six cases (23.1 %), pulmonary bleeding in two cases (7.7 %), and arterial thrombosis or thromboembolism also in two cases (7.7 %). In addition, we found individual cases with invasive pulmonary mycosis or gastric peptic ulcer as conditions leading to cause of death (3.8 % each). Of note, in three cases (11.5 %) we did not find pathologies that would qualify for an intermediate between underlying and immediate cause of death.

Next, we identified underlying causes of death, i.e. diseases that initiated the events resulting in death (Table 2). For this, we considered all autopsy findings and the clinical history to determine the underlying disease that would causally explain conditions leading to cause of death and immediate causes of death in all cases. We determined COVID-19 pneumonia as underlying cause of death in the majority of decedents (53.8%). Of note, when considering COVID-19 or SARS-CoV-2 infection as underlying cause of death or condition leading to cause of death, this applied to a total of 24 cases (92.3 %). Preexisting lung emphysema, pulmonary hypertension, and chronic right ventricular insufficiency were considered as diseases underlying cause of death in 26.9 %. In two cases (7.7 %), cardiovascular disease, and in two further cases (7.7 %), malignant tumors or consequences of tumor therapy were considered as underlying disease. Collectively, our findings demonstrate that septic organ failure, pneumonia, respiratory insufficiency, and right ventricular heart failure due to COVID-19 were the most frequent pathological mechanisms of death in these patients.

### Comorbidities found by autopsy in COVID-19 decedents

To learn about other relevant disease conditions in patients that died after SARS-CoV-2 infection, we determined the presence and extent of comorbidities by autopsy (Table 2). Generalized atherosclerosis was present in all but one case (96.2%), and was mild in 9 (34.6 %), moderate in 3 (11.5 %) and severe in 13 cases (50 %). Similarly, coronary artery disease was seen in all but two cases (92.3 %), and was mild in 8 (30.8 %), moderate in 4 (15.4 %) and severe in 11 decedents (42.3 %). Furthermore, we noticed preexisting emphysema of the lungs in almost half of the cases (46.2 %), and pulmonary artery sclerosis as a sign of pulmonary hypertension in 11 cases (42.3 %). Other comorbidities included myocardial hypertrophy (65.4%), hepatosteatosis (11.5%) and liver fibrosis (7.7%). In two cases (7.7 %), we defined COVID-19 or SARS-CoV-2 as comorbidity, since we did not observe a direct contribution of this infection to the mechanism of death. These findings demonstrated a high prevalence of cardiovascular and pulmonary comorbidities in patients that had died after COVID-19.

## DISCUSSION

Here, we present data on causes of death and comorbidities in patients that had died after a severe course of COVID-19. These patients had reached a median age of 70 years, which in line with previous reports indicates increased risks for fatal COVID-19 outcome with older age^12,13^. Clinical records showed that in the majority of cases, respiratory insufficiency was a dominating symptom, while the most frequent clinical cause of death was sepsis and thus infection related. In line with these findings, we found by autopsy that sepsis caused by purulent bacterial lung infection was the most frequent cause of death, while, in some cases, we observed deadly respiratory insufficiency due to diffuse alveolar damage. These findings suggested that SARS-CoV-2 infection could directly cause lethal lung damage. However, death due to pulmonary bacterial superinfection and sepsis appeared to be a more common causal chain of events that may significantly endanger patients with severe COVID-19-related lung damage. We hypothesize that such causality may be even more prevalent in clinical settings where respiratory insufficiency is manageable by mechanical ventilation or extracorporeal oxygenation.

A recent study showed high frequencies of fatal pulmonary thromboembolism in patients that had died of COVID-19^10^. Deep venous thrombosis was identified as the likely thromboembolic source. In line with these findings, pulmonary thromboembolism was diagnosed in almost every fourth of our patients during the clinical course of the disease. However, we found that pulmonary thromboembolism was an immediate cause of death or a condition leading to cause of death in two cases only, which suggests that in other cases hypercoagulability may have been effectively controlled by anti-coagulant treatment. Nevertheless, hypercoagulability appears to be an important, severe and potentially fatal aspect of COVID-19^14,15^. Furthermore, peripheral microthrombosis in multiple organ systems has been reported^16^, which may cause severe organ damage also in patients that survive COVID-19. It therefore remains to be determined by prospective clinical studies, if anti-coagulation reduces the risk of COVID-19 related death, and to what extent this affects the risk of organ damage in COVID-19 survivors.

The majority of deceased patients in our study had diagnosed comorbidities that with arterial hypertension, chronic kidney or heart disease, and chronic pulmonary disease most frequently affected the cardiovascular and respiratory system. By autopsy, we confirmed these clinical diagnoses, since we found their pathological correlates that included general and coronary atherosclerosis, cardiac hypertrophy, and pulmonary emphysema amongst others. In addition, we found a high prevalence of lifestyle risk factors, such as obesity, alcohol consumption, and nicotine abuse. These findings are in agreement with previous studies and imply that patients with preexisting chronic health conditions or lifestyle risk factors are at an increased risk for fatal outcome of COVID-19^17,18^. However, considering both the high frequency of these comorbidities and the relatively high age of patients that died after SARS-CoV-2 infection, this led to a reasonable debate about the extent to which preexisting health conditions or COVID-19 determined the time of death^19^, and our data may further inform about this issue. We found that sepsis due to lung infections and respiratory insufficiency were the most frequent immediate causes of death. However, our autopsy series included no single case of immediate deadly ischemic heart disease or stroke, which are the most common causes of death worldwide^20^, and for which the majority of comorbidities as well as the mentioned life style risk factors that we found were strongly predisposing^21^. This indicates that the immediate causes of death appear not to be caused directly from the observed comorbidities but instead were directly linked to lung damage initiated by SARS-CoV-2 infection in most cases. Our data therefore suggest that in the majority of our cases with severe and fatal COVID-19, patients had died of this disease, although in the presence of multiple preexisting health conditions. These findings also support the idea that patients who died of COVID-19 appear to have lost considerable lifetime, independent of their age, as reported by others^19^.

A limitation of our study is the relatively small sample size. Furthermore, patients included in this study had reached a median age of 70 years, which mirrors reported age distributions of inpatient non-survivors in Wuhan^12^, and data from a recent autopsy report^16^, but is lower than suggested by other epidemiologic data from Italy on COVID-19 decedents^22^. While regional factors may influence age distribution, this discrepancy also suggests a case selection bias, and we speculate this may reflect which patients were hospitalized and therefore received most intense therapeutic measures. The interpretation of autopsy results and conclusions on health impacts of COVID-19 therefore requires careful consideration of the study population.

## Data Availability

All relevant information is included in the manuscript. Additonal data is available from the correspondig author upon reasonable request.

## ACKNOWLEDGEMENTS

We are indebted to Anistan Sebastiampillai, Juliane Plaschke and Francisca Egelhofer for excellent technical assistance and helpful advice. The authors declare no conflicts of interest.

## AUTHORS CONTRIBUTIONS

S.E., S.G., J.I., E.M., P.B., C.K., B.I.H., H.H., H.M., H.R., F.H. and D.H. performed autopsies, histopathology and clinical workup. V.M.C. made viral RT-qPCR workups. D.H. supervised the study. All authors analyzed the data, wrote, revised and approved the manuscript.

